# Reassessing Instrument Strength in Two-Sample Mendelian Randomization Analysis

**DOI:** 10.64898/2026.06.16.26355811

**Authors:** Xiaonan Liu, Yu-Jyun Huang, Yogesh Purushotham, Tamar Sofer

## Abstract

Mendelian randomization (MR) analysis is widely used to estimate causal relationships between risk factors and outcomes of interest. Two-sample MR approaches have gained increasing attention in genetic epidemiology due to the growing availability of Genome-Wide Association Study (GWAS) summary statistics from public databases. A critical step in two-sample MR is the selection of genetic variants as instrumental variables (IVs). Although genome-wide significant variants are typically preferred, the inclusion of variants with weaker association p-values is considered, as they may potentially improve power through an increased instrument number of instruments, while they may introduce weak instrument bias and attenuate effect estimates towards the null. Our simulation results show that even modest levels of pleiotropy substantially increase the variability of causal effect estimates, while the inclusion of weak IVs does not substantially affect the direction and variability of causal effect estimates in most cases. In real data analyses, we used two released versions of FinnGen GWAS summary statistics with different sample sizes as exposure GWASs to assess the influence of weak IVs. Here, the inclusion of IVs with higher exposure-association p-values resulted in weakened estimated effect sizes, particularly when the exposure GWAS sample size was small. These findings suggest that incorporating weak IVs is reasonable when the exposure GWAS sample size is large, but it poses a risk of falsely concluding null associations when the exposure GWAS sample size is small.

## Introduction

Mendelian randomization (MR) is a widely used approach for estimating the causal effects of exposures on outcomes using genetic variants as instrumental variables (IVs) (Davey Smith and Hemani 2014; Zheng et al. 2017; Chen et al. 2024). The increasing availability of publicly shared Genome-Wide Association Study (GWAS) summary statistics enables researchers to perform two-sample MR using data from independent GWASs, without requiring access to individual-level data (Slob and Burgess 2020; Sanderson et al. 2022; Hu et al. 2024). Valid MR inference relies on three core assumptions: (i) the genetic instruments are associated with the exposure (relevance); (ii) the instruments are independent of confounders of the exposure-outcome relationship (independence); and (iii) the instruments impact the outcome only through the exposure (exclusion restriction). In practice, inference of MR also relies on the strength of instruments, characterized by the strengths of association with the exposure. Consequently, numerous methodological advances have been developed to evaluate and address violations of key assumptions in the MR analysis, including approaches for detecting pleiotropy (Verbanck et al. 2018; Morrison et al. 2020) and reducing bias due to weak instruments (Zhao et al. 2020; Wang and Kang 2022). Although the seminal paper introducing the two-sample MR framework recommended a liberal p-value threshold of 10^−5^for IV selection (Burgess, Butterworth, and Thompson 2013), most studies employ the more stringent genome-wide significance threshold (i.e., p-value < 5 × 10^−8^), both to limit weak-instrument bias and to reduce the cumulative risk of assumption violations that grows with the number of included IVs in the analysis.

While this strategy is standard, it may limit applicability in certain contexts, for example, GWASs conducted in underrepresented populations or those affected by participation bias due to structural or demographic factors (e.g., the Million Veteran Program, in which only around 10% of participants are female (Huang et al. 2025)). Furthermore, the limited number of studies investigating the impact of incorporating varying numbers of genetic instruments, whether by relaxing the p-value threshold or, in more extreme cases, relying solely on weak IVs, has hindered the broader application of MR for populations or exposures with small GWAS sample sizes.

Motivated by this gap, this study aims to empirically evaluate how different instrument selection thresholds influence causal effect estimates within the two-sample MR framework. To study this, we performed extensive simulation studies, as well as real-data analyses. In the simulation studies, we assessed the impact of varying the p-value threshold for IV selection, while also varying the exposure GWAS sample size, exposure trait heritability, degree of pleiotropy, and number of variants causal to the exposure. In real data analysis, we assessed the impact of relaxing the p-value threshold to include more IVs, in settings of low and high exposure GWAS sample sizes. In data analysis, we considered the impact of weak IV inclusion in two ways: (i) including weak instruments in addition to strong instruments, and, in secondary analysis, (ii) extreme scenarios assuming that only weak instruments are available. We estimated the causal effects of six cardiovascular risk factors using GWAS summary statistics from FinnGen (Kurki et al. 2023) and four aging-related chronic disease outcomes with summary statistics obtained from the Pan-UK Biobank European ancestry individuals (Karczewski et al. 2025). Our results differ between simulations and data analyses. In simulations, the results are overall similar with and without inclusion of IVs with higher association p-values. In data analysis, including additional weak IVs often attenuates effect estimates towards the null, with this phenomenon being more pronounced when the exposure GWAS sample size is low. This discrepancy between simulation and data analysis results suggests that the impact of weak IVs in real-world MR analyses may extend beyond classical weak instrument bias alone. In particular, relaxing IV selection thresholds may increase the inclusion of invalid instruments that are more likely to violate core MR assumptions through horizontal pleiotropy or residual confounding. Although the inclusion of weakly associated variants may have limited impact when the exposure GWAS is sufficiently powered, our analyses suggest that greater caution, including additional sensitivity analyses, is warranted when applying more liberal IV selection thresholds in settings with smaller exposure GWAS sample sizes.

## Methods

### Simulation studies

We conducted simulation studies to systematically evaluate the impact of p-value thresholds for IV selection and consequent MR-based estimation under controlled settings. The simulation framework was designed for varying key parameters, including the number of causal variants, sample size, and degree of pleiotropy. We used the sim1000G R package (Dimitromanolakis et al. 2019) to simulate genotype data for *N* = 30,000 individuals using the phased European population of the 1000 Genomes Project as a reference panel. Variants with minor allele frequency (MAF) below 1% were excluded prior to simulation. Variants were simulated in groups of 100 independent linkage disequilibrium (LD) blocks, yielding a total of *p* = 501,378 variants across blocks, including 343,175 variants with MAF ≥ 0.05. These LD blocks were selected from a list of blocks defined in (Berisa and Pickrell 2016). We generated a continuous exposure *X* using the following equation:

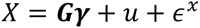

where ***G*** denotes the *N* × *p*_*causal*_ genotype matrix of randomly selected causal common variants (MAF ≥ 0.05) and γ being the *p*_*causal*_ × 1 vector of their effect sizes, with *p*_*causal*_ ∈ {10, 100} and each variant was chosen from a different LD block. The genetic effect size of SNP *i* was sampled as γ_*i*_∼ *N*(0, ℎ^2^/*p*_*causal*_) then scaled by allele frequency by dividing 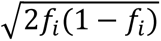 where ℎ^2^ ∈ {0.4, 0.6} represents the exposure heritability, *f*_*i*_ represents the allele frequency of SNP *i*. The unobserved confounder was generated as 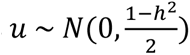, and the random error term as 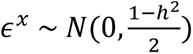.

We generated a continuous outcome *Y* using the following:

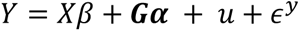

where the true causal effect was fixed at *β* = 0.1, ***α*** represents pleiotropic effects with 70% of variants having α_*i*_ = 0, and the remaining 30% of causal variants (*p*_*nonzero*_) had pleiotropic effects sampled from *N*(0,0.4/*p*_*nonzero*_), followed by the same adjustment for allele frequency as above, and *ϵ*^*y*^ ∼ *N*(0,1) denotes the independent error term. For comparison, we also simulated a scenario with no pleiotropy, where ***α*** = **0**.

We simulated exposure and outcome for all 30,000 individuals. To mimic a two-sample MR setting, we split the samples into: *N*_*exposure*_ ∈ {10000, 20000} individuals for the exposure GWAS and 10,000 individuals for the outcome GWAS. We first conducted an “oracle” MR analysis where the true causal variants were assumed to be known and were directly used as instrumental variables in the MR analysis. This provides a baseline of MR estimates when all IVs are causal to the exposure (though some of the IVs may be pleiotropic and therefore not valid). In the non-oracle MR analysis causal variants were assumed to be unknown and were selected based on applying different p-value thresholds based on the exposure GWAS, and clumping. For both settings, we estimated the causal effect using two-sample MR methods based on summary statistics obtained from the exposure and outcome GWAS. We repeated the simulations 100 times for each scenario (defined by *N*_*exposure*_, *p*_*causal*_, ℎ^2^, ***α***) and computed the estimated causal effects and coverage across 100 simulations.

### Data analysis

We selected common diseases with established GWAS as outcomes and known risk factors as exposures. Outcomes included atrial fibrillation (AF), coronary artery disease (CAD), stroke, and Alzheimer’s disease (Alz). Exposures included type 2 diabetes (T2D), hypertension (HTN), obesity, sleep disorder, chronic kidney disease (CKD) and disorders of lipoprotein metabolism and other lipidaemias (LpA). GWAS summary statistics for exposures were obtained from FinnGen Releases 3 (135,638 individuals) and 12 (∼480,000 individuals), for low and high exposure GWAS sample size settings. All of the FinnGen participants are of Finnish ancestry. To ensure non-overlapping study cohorts from the exposure GWAS, the GWAS for all outcomes except Alz were obtained from the Pan-UK Biobank (UKB) project, restricted to the European ancestry individuals. Although there are differences between non-Finnish, British European, and Finnish genetic ancestries, genetic associations often replicate between them (Sun et al. 2022), and effect sizes of common, highly associated variants, are highly correlated between the Finnish and the UKB biobanks (Kurki et al. 2023). Due to the limited number of Alz cases from Pan-UKB, we used GWAS results from (Schwartzentruber et al. 2021), a meta-analysis of the (Kunkle et al. 2019) GWAS of diagnosed Alz and GWAS for family history of Alz in the UKB. Table 1 summarizes information from GWAS used in our analysis.

**Table 1.** Summary of exposure and outcome GWASs used in the MR analysis. Overview of GWASs used for exposures and outcomes. Selected exposures are shown in the upper section of the table, and outcomes are listed in the lower section. The second column lists the consortium or study from which the GWAS summary statistics were obtained. The third column summarizes the number of variants reaching genome-wide significance (p-value < 5 × 10^−8^). The fourth column reports the total sample size of the original GWAS, along with the number of cases for each trait. Note: Alz here includes diagnosed late-onset Alz or a family history of Alz. Abbreviation: GWAS: genome-wide association study; LpA: lipoprotein metabolism and other lipidaemias; CKD: chronic kidney disease; HTN: hypertension; T2D: type 2 diabetes; AF: atrial fibrillation; Alz: Alzheimer’s disease; CAD: coronary artery disease; UKB: UK Biobank; IGAP: international genomics of Alzheimer’s project.

| Phenotype | Consortium | Number of genetic variants at genome-wide association threshold | Total sample size (cases) |
| --- | --- | --- | --- |
| **Exposures** |  |  |  |
| LpA | FinnGen | 8,131 | 479,432 (59,366) |
| CKD | FinnGen | 2,166 | 493,235 (12,787) |
| Sleep disorder | FinnGen | 2,008 | 495,270 (65,076) |
| Obesity | FinnGen | 3,651 | 500,192 (31,499) |
| HTN | FinnGen | 29,707 | 500,264 (154,630) |
| T2D | FinnGen | 22,087 | 486,367 (82,878) |
| **Outcomes** |  |  |  |
| AF | UKB | 3,311 | 420,531 (19,218) |
| Alz | UKB and the IGAP | 3,859 | UKB: 431,000 (53,000)<br>IGAP: 64,000 (22,000) |
| CAD | UKB | 3,420 | 420,531 (31,148) |
| Stroke | UKB | 2 | 419,624 (6,454) |

GWAS summary statistics were processed using the following steps: (i) in the exposure GWAS, variants with duplicate rsIDs or MAF < 0.01 were removed; (ii) genomic coordinates of the exposure GWAS from FinnGen were converted from GRCh38 to GRCh37 using the UCSC LiftOver tool (Hinrichs et al. 2006) to match the reference build of the Pan-UK Biobank; (iii) exposure and outcome summary statistics were merged (for each exposure-outcome combination); (iv) variants were selected based on GWAS p-value thresholds of 10^−5^, 10^−6^, 10^−7^, and 5 × 10^−8^; (v) LD clumping was performed using the European reference panel from the 1000 Genomes Project, with clumping parameter R^2^ = 0.001 and distance = 10 kb. Finally, exposure and outcome summary statistics for each pair were harmonized to align effect alleles and remove ambiguous variants.

We conducted two-sample MR methods using variants obtained using each p-value thresholding. Methods included weighted median (Bowden et al. 2016), inverse variance weighted meta-analysis (IVW) (Burgess, Butterworth, and Thompson 2013), penalized robust IVW (Rees et al. 2019), MR-Egger (Bowden, Davey Smith, and Burgess 2015), penalized robust MR-Egger (Rees et al. 2019), MR-PRESSO (Verbanck et al. 2018), MR-ConMix (Burgess et al. 2020), and MR-RAPS (Zhao et al. 2020). Primary analyses were conducted using MR-RAPS, a statistically rigorous approach that corrects weak instrument bias by accounting for measurement error in SNP-exposure associations via an adjusted profile likelihood and employing robust loss functions to down-weight pleiotropic outliers, enabling consistent and computationally efficient estimation with many weak instruments. When four or fewer variants were available, IVW was used instead. To further assess the impact of using weak instruments only, in a secondary analysis, we grouped variants based on exposure p-values into the following intervals: [0, 5 × 10^−8^), [5 × 10^−8^, 10^−7^), [10^−7^, 10^−6^), and [10^−6^, 10^−5^] and performed separate clumping for each interval. In this analysis, the IVs used for MR analysis were non-overlapping across intervals. Finally, we also performed multivariate MR (MVMR) analysis including obesity as an exposure in addition to T2D, HTN, LpA, CKD, and sleep disorders, to assess the impact of varying IV inclusion p-value threshold in the MVMR context. Analyses were run in R version 4.3.2 (TwoSampleMR (Hemani et al. 2018) for MVMR analysis and MendelianRandomization R packages (Yavorska and Burgess 2017) for two-sample MR analyses). Throughout this paper, we use p-values for two distinct purposes: (i) p-value thresholds for selecting IVs, which vary across analyses and are specified where relevant; and (ii) the p-value of the causal effect estimate, which we compare against α = 0.05 to determine statistical significance of the causal inference.

## Results

### Simulation studies

Figure 1 summarizes the performance of oracle and non-oracle analyses using MR-RAPS across 100 simulations under ℎ^2^ = 0.4, *p*_*causal*_ = 100, *N*_*exposure*_ = 20,000 and 10,000 with pleiotropy. The upper panel shows the distribution of causal effect estimates and the lower panel shows the corresponding mean squared error (MSE) across the simulations. Across both exposure GWAS sample sizes, the oracle estimates were centered around the true effect and had low MSE across models, providing validation of our simulation setup. In the non-oracle analysis, estimates were similar across p-value thresholds, with low dispersion and MSE, indicating low bias and variance of estimates. While performance was generally similar across p-value thresholds, we observed more apparent differences across MR methods. Penalized robust MR-Egger, MR-Egger, and IVW showed greater variability in their causal estimates and higher MSE than other methods, suggesting less stable performance under this simulation setting.

**Figure 1.**
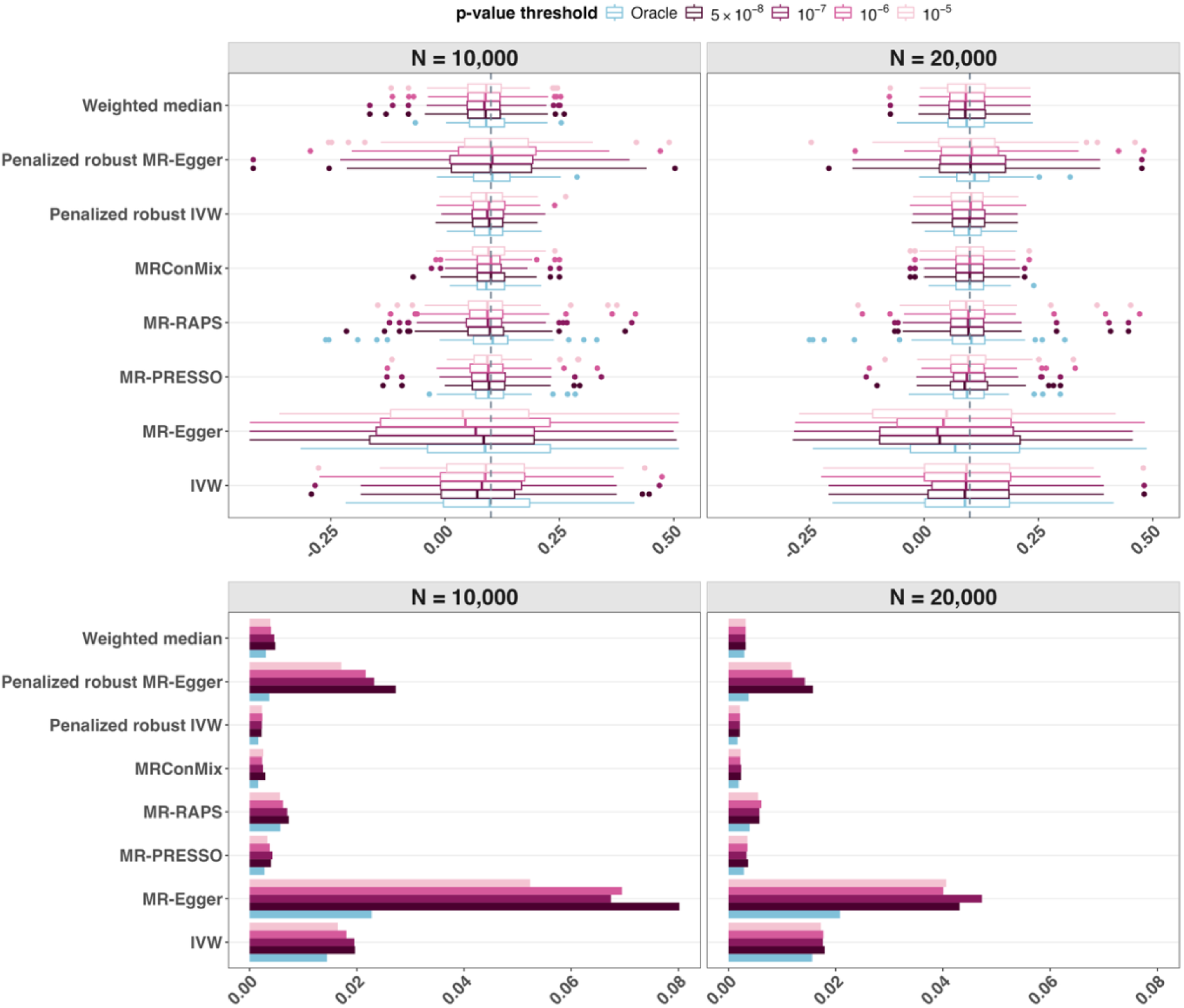
Simulation performance of oracle and non-oracle analyses under ℎ^2^ = 0.4, *p*_*causal*_ = 100, *N*_*exposure*_ = 20000, 10000 with pleiotropy. (a) Boxplot of MR causal effect estimates across 100 simulations. Oracle MR (blue) uses the true causal variants as instruments; non-oracle MR (pink shades) selects instruments from the exposure GWAS using different p-value thresholds. Grey dashed line indicates true causal effect. (b) Mean squared error (MSE) computed across 100 simulations.

When comparing the two exposure GWAS sample sizes, the overall patterns were similar between *N*_*exposure*_ = 20,000 and 10,000. Additional simulation settings (defined by *N*_*exposure*_, *p*_*causal*_, ℎ^2^, with and without pleiotropy) are shown in Supplementary Figures 1-7 and Supplementary Table 1. Across all settings, the general performance patterns are consistent. The most notable difference is observed under pleiotropy, compared to no pleiotropy, where estimates show greater variability and MSE values are higher, especially in the setting of 10 causal variants.

### Data analysis

Figure 2 shows the estimated causal associations between exposures and outcomes across a range of p-value thresholds applied over the exposure GWAS summary statistics. We report results from MR-RAPS in the primary analysis due to its robustness against weak instruments. In general, as our exposures are known CVD risk factors, the estimated exposure-outcome causal associations tended to be risk increasing, as expected, for the three CVD outcomes (AF, stroke, and CAD). In contrast, the associations were often protective for Alz, other than sleep disorders, for which the estimated associations were usually null or risk increasing, though mostly not statistically significant.

**Figure 2.**
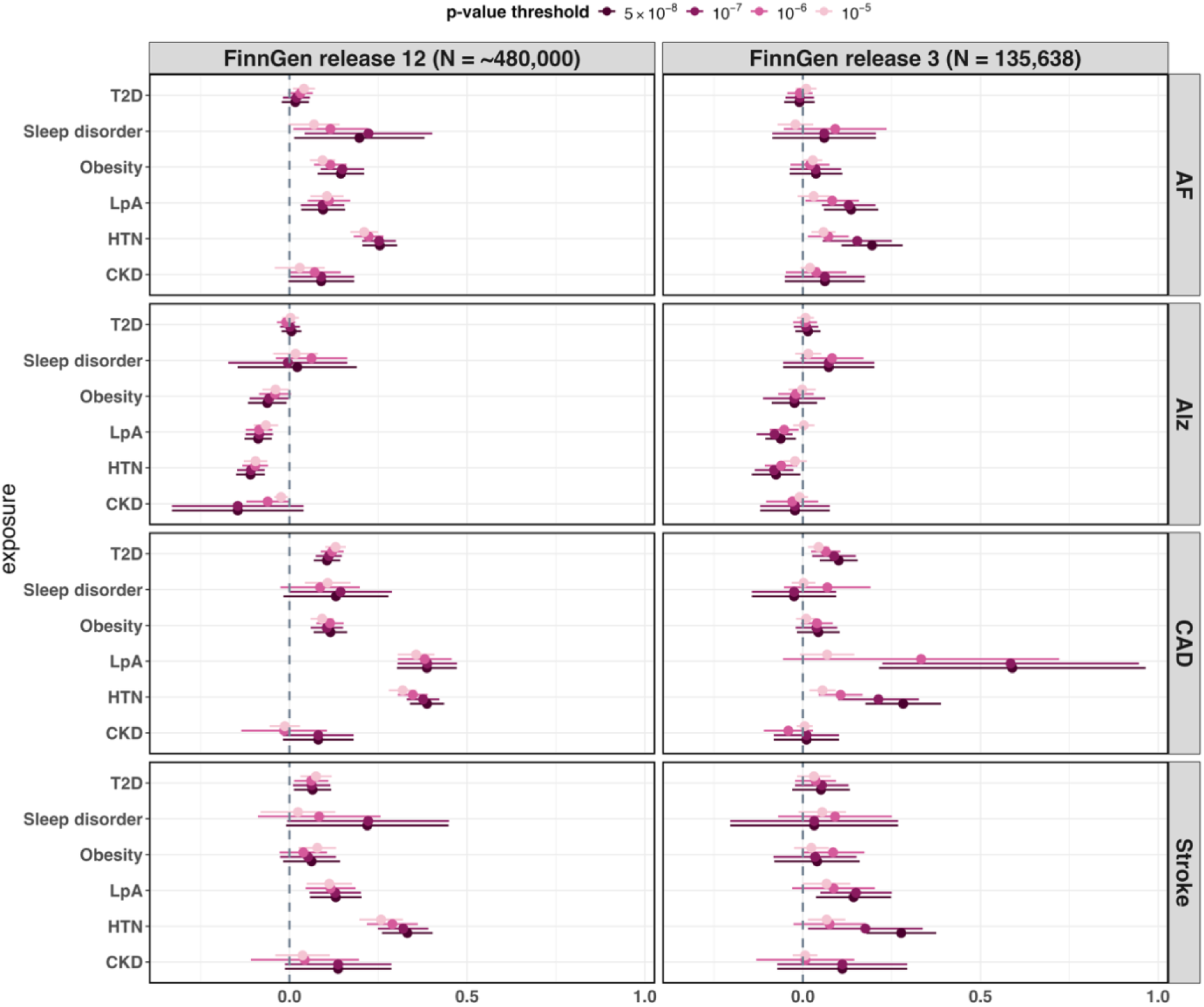
Forest plot showing causal effect estimates (log odds scale) and corresponding 95% confidence intervals for each exposure-outcome pair obtained using MR-RAPS. Exposures are listed on the y-axis of each panel, and outcomes are shown on the right panel. The left panel shows the results where instrumental variables were selected using different p-value thresholds from FinnGen release 12 and the right panel shows the results using a smaller sample size of exposure GWAS (FinnGen release 3). The vertical dotted line indicates the null causal effect. Abbreviation: AF: atrial fibrillation; Alz: Alzheimer’s disease; CAD: coronary artery disease; LpA: lipoprotein metabolism and other lipidaemias; CKD: chronic kidney disease; HTN: hypertension; T2D: type 2 diabetes

Across p-value thresholds, the direction (sign) of effect estimates was consistent across thresholds for most exposure-outcome pairs. Similarly, association testing results at the α = 0.05 level were mostly consistent across thresholds. In FinnGen release 12, when comparing the genome-wide significance threshold 5 × 10^−8^ with the relaxed threshold 10^−5^, 23 of 24 exposure-outcome pairs had effect sizes in the same direction, and 19 pairs led to the same testing results at the α = 0.05 level. For instance, HTN was consistently associated with increased risk for all outcomes, obesity was consistently (i.e. across p-value thresholds) associated with increased risk for AF and CAD, and T2D was consistently associated with increased risk for CAD and stroke.

To assess whether the impact of p-value thresholds differed in a smaller exposure GWAS dataset, we repeated the analyses using FinnGen release 3. The results showed slightly less consistency across thresholds: 20 of the 24 pairs had the estimates in the same direction when comparing 5 × 10^−8^ with 10^−5^, and 18 pairs had the same causal effect testing results at the α = 0.05 level. Notably, the attenuation of causal effect estimates toward the null was more pronounced than in release 12, which has a larger sample size. In several exposure-outcome pairs, such as HTN on AF and HTN on CAD, the confidence intervals of causal estimates using a p-value threshold of 10^−5^ were narrow and shifted close to the null, with little or no overlap with those obtained using a 5 × 10^−8^ threshold for IV selection, even though the causal effect testing conclusions were consistent at α = 0.05. Nevertheless, the overall pattern remained similar to FinnGen release 12, suggesting that the exposure GWAS p-value threshold had limited impact on the direction and the causal effect testing conclusions at α = 0.05 level, although effect-size estimates themselves can be attenuated when the exposure GWAS sample size is smaller. Similar patterns were observed when using the standard IVW method (Supplementary Figure 8). Complete results are provided in Supplementary Table 2.

When comparing the results with non-overlapping p-value intervals, most conclusions were the same, except for the associations between pairs of LpA-Alz and CKD-CAD using p-value interval [5 × 10^−8^, 10^−7^) and CKD-Stroke using p-value interval [10^−7^, 10^−6^) where the direction of estimates changed (Supplementary Figure 9-10). In the overlapping-threshold analysis, 10^−5^showed the tightest confidence intervals, likely because it included the largest number of variants. In contrast, in the non-overlapping p-value interval analysis, estimates varied more across p-value intervals and confidence intervals were wider, reflecting fewer variants within each interval. This variability was more evident when using FinnGen release 3 with a smaller sample size.

Interestingly, including weaker instruments resulted in a stronger estimated association for T2D on AF, which became statistically significant (null estimates at p-value thresholds 5x10^-8^ to 10^-6^, but estimated causal OR=1.04, 95% CI (1.01, 1.07) when using p-value threshold 10^-5^). This estimate remained statistically significant when using MR-PRESSO. Indeed, looking at MR using only weak IVs, here 461 variants with 10^-6^<p-value<10^-5^, the association was statistically significant. Thus, in this case, the weak instruments alone suggest a causal association between T2D and AF.

To assess potential violations of the no horizontal pleiotropy assumption when adding variants as IVs, we compared MR-PRESSO and MR-RAPS results (Figure 3), which were consistent across methods. Similar patterns were observed using non-overlapping p-value intervals (Supplementary Figure 11), suggesting no evidence of increased horizontal pleiotropy when increasing p-value thresholds. Lastly, we performed MVMR analysis (Supplementary Figure 12), to adjust for obesity, as a CVD risk factor that is also associated with other exposures, and therefore may confound the exposure-outcome causal effect. Most conclusions remained consistent except for the association between LpA and Alz, where the estimate changed direction from being protective (MR) to risk increasing (MVMR).

**Figure 3.**
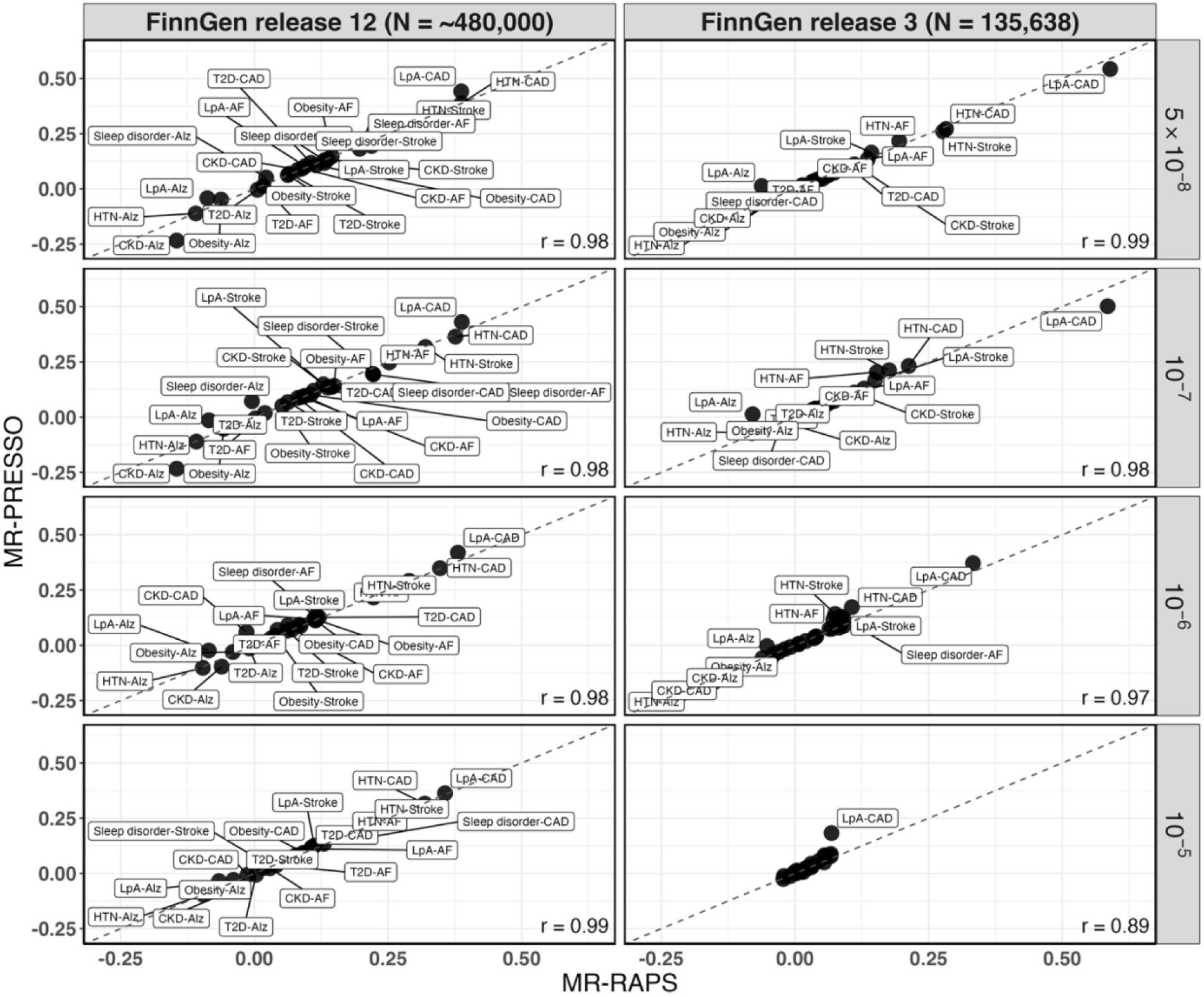
Comparison of estimated causal effects from MR-PRESSO (y-axis) and MR-RAPS (x-axis) using p-value thresholds. Results using FinnGen release 12 are shown on the left, while those based on FinnGen release 3 are shown on the right. Abbreviation: AF: atrial fibrillation; Alz: Alzheimer’s disease; CAD: coronary artery disease; LpA: lipoprotein metabolism and other lipidaemias; CKD: chronic kidney disease; HTN: hypertension; T2D: type 2 diabetes. The bottom right corner shows the Pearson’s correlation.

## Conclusion

This study evaluates the impact of using more liberal p-value thresholds, such as p-value < 10^−5^, for IV selection in two-sample MR analyses. While simulation studies varying key parameters resulted in relatively homogeneous findings, data analysis results were much more sensitive to the sample size of the exposure GWAS, and to the p-value threshold for IV selection.

While patterns of causal effect estimates across p-value thresholds and exposure GWAS sample sizes vary between exposure-outcome pairs, we conclude that including weaker instruments is risky when the exposure sample size is small, as causal effect estimates are attenuated towards the null while confidence intervals are small. Critically, these confidence intervals often do not cover the causal effect estimates obtained from MR performed with a large sample size exposure GWAS and genome-wide significant variants as instruments. Thus, the risk is of wrongly concluding a weak or null causal effect. Still, as MR estimates often lack specific interpretation, it may still be useful to use weak instruments and focus on likelihood of existence and the direction of causal effects when interpreting results. Comparing causal estimates across different MR approaches, which are developed to account for different assumptions, is another useful way to validate these interpretations.

Our simulation studies showed that relaxing the p-value thresholds generally produced similar causal estimates across a range of simulation settings. In the empirical data analyses, relaxed thresholds mostly led to the same directions of estimates and testing results but the patterns of estimates showed greater variation than in simulations. One possible explanation is that real-world GWAS contain complexities that are difficult to capture or design in simulation studies, including ill-defined exposure (e.g., “sleep disorder” is likely a combination of multiple potential sleep problems, and these problems may have different genetic determinants and different relationships with the outcome of interest), and, more generally, complicated causal structures where genetic variants associated with the exposure are also associated with unobserved confounders of the exposure-outcome relationship. These factors may explain why p-value thresholds have a greater impact on MR results in real data analyses than in our controlled simulation studies.

Based on our simulation and empirical investigations, we suggest that more liberal p-value thresholds may be considered for IV selection in MR analyses. This recommendation is consistent with the view that MR estimates should not be over-interpreted as precise causal effect sizes, but rather as evidence supporting or rejecting a causal hypothesis (Woolf and Burgess 2025). From this perspective, some variation in point estimates across p-value thresholds may be acceptable as long as the direction of effect and the conclusion of statistical inference of causal effects remain consistent. However, the impact of relaxing the p-value threshold appeared to be more pronounced when the exposure GWAS sample size was smaller. We observed that estimates tend to show greater variability and attenuation towards the null, even for strong associations such as HTN on AF. Therefore, when the exposure GWAS sample size is small, causal effect estimates and their statistical inference conclusions should be accompanied by sensitivity analyses across multiple p-value thresholds to assess the robustness of the findings. Along with recently proposed robust MR methods, this approach may reduce the power limitations of IV selection while maintaining the validity of causal inference. We anticipate that this strategy could broaden the applicability of two-sample MR, particularly for exposures with underpowered GWAS data.

## Data Availability

This study used publicly available GWAS summary statistics from the FinnGen study (https://www.finngen.fi/en/access_results) and Pan-UK Biobank project (https://pan.ukbb.broadinstitute.org/).

## Acknowledgements

This work was supported by the U.S. National Institute on Aging grant R01AG080598. This study used publicly available GWAS summary statistics from the FinnGen study (https://www.finngen.fi/en/access_results) and Pan-UK Biobank project (https://pan.ukbb.broadinstitute.org/). We acknowledge the participants and investigators of the FinnGen study. We also acknowledge the Pan-UK Biobank project and the UK Biobank resource under Application Number 31063.

## Notes

### Competing Interest Statement

The authors have declared no competing interest.

